# Impact of Prehospital Triage Protocol on Outcomes of Spontaneous Intracerebral Hemorrhage Patients

**DOI:** 10.1101/2025.05.30.25328685

**Authors:** Tracey H Fan, Molly Lawrence, Elena Goicoechea, Angela Wick, Shyam Prabhakaran

## Abstract

**Background:** While prehospital triage protocols for suspected large vessel occlusion (LVO) improve ischemic stroke outcomes, their impact on spontaneous intracerebral hemorrhage (sICH) remains uncertain. We evaluated whether a regional LVO-focused emergency medical service (EMS) transport protocol affected care efficiency and outcomes in sICH patients.

**Methods:** We conducted a multicenter pre-post implementation cohort study using the Get-With-The-Guidelines-Stroke database in Chicago (April 2017–January 2020).

Included were EMS-transported sICH patients arriving ≤6 hours from last known normal at 8 comprehensive stroke centers (CSCs) and 15 primary stroke centers (PSCs). Primary outcomes were in-hospital mortality and favorable discharge disposition (home/acute rehabilitation). Secondary outcomes included good neurologic outcome (independent ambulation) at discharge and time metrics (symptom-to-arrival, door-to-CT). Interrupted time series (ITS) analysis assessed changes while accounting for temporal trends.

**Results:** Among 311 sICH patients (111 pre-, 192 post-implementation), there was no difference in in-hospital mortality (12% vs. 9%, p=0.4; ITS level change: −5% [95% CI: −31% to 21%], p=0.68; trend change: 1% [95% CI: −1% to 2%], p=0.34), favorable discharge disposition (58% vs. 64%, p=0.3; ITS level change: −20% [−77% to 38%], p=0.49; trend change: 1% [95% CI: −2% to 4%], p=0.47) or good neurologic outcomes (13% vs. 19%, p=0.4; ITS level change: 11% [−25% to 48%], p=0.53; trend change: −1% [95% CI: −3% to 1%], p=0.37) between pre-post implementation periods. Time metrics (door-to-CT, symptom-to-arrival, symptom-to-CT) showed no significant changes in unadjusted or ITS analyses. The protocol also did not impact CSC admissions rate and inter-hospital transfers in ITS analyses.

**Conclusion:** Implementation of an LVO-focused EMS transport protocol did not improve outcomes or care efficiency among sICH patients, nor did it affect CSC admission or transfer rates. These findings highlight the need for dedicated prehospital triage strategies specific to sICH, distinct from ischemic stroke pathways.

## Introduction

Spontaneous intracerebral hemorrhage (sICH) represents approximately 10-15% of all strokes, with incidence increasing in recent years with more widespread use of anticoagulants.^1,2^ sICH is also associated with the highest morbidity and mortality among stroke subtypes with early-term mortality reaching 30-40%.^1^ Timely hospital arrival, diagnostic imaging, and intervention are critical in acute stroke management, as delays have been consistently linked to worse neurological outcomes and increased mortality.^3^ Unlike ischemic stroke, where reperfusion therapies offer clear and time-sensitive intervention, sICH care is focused on blood pressure control, anticoagulation reversal, and early neurosurgical intervention (e.g., external ventricular drainage, minimally invasive surgery [MIS]) when indicated.^4–6^

Prehospital triage protocols that transport patients with suspected large vessel occlusion (LVO) to the comprehensive stroke center (CSC) have suggested benefits such as reduced time to EVT, increased rates of EVT and improved functional outcomes for ischemic stroke patients in urban settings.^6–8^ A secondary analysis of the RACECAT trial found that sICH patients transported directly to CSCs experienced worse 90-day functional outcomes, likely due to longer transport times without corresponding benefits from specialized interventions.^9^ However, the benefit of these triage protocols in sICH patients in more organized, urban settings remains unclear. In this study, we aimed to evaluate whether a regional prehospital transport protocol specifically designed for suspected LVO patients also impacted care efficiency, in-hospital mortality, and early neurological outcomes among sICH patients in Chicago, Illinois.

## Methods

We conducted a multi-center pre-post implementation cohort study using the Get-With-The-Guideline-stroke database (GWTG-stroke; IQVIA, Inc) of consecutive patients at Chicago’s stroke centers; data were analyzed from April 1, 2017 until January 31, 2020. This preimplementation-postimplementation study was approved by the institutional review board at the University of Chicago, Chicago, Illinois. Participating sites were granted a waiver of informed consent under the common rule because the data are collected from medical records for local quality improvement purposes and aggregated in a deidentified manner for regional reporting. The study is reported according to the Strengthening the Reporting of Observational Studies in Epidemiology (STROBE)^10^ reporting guidelines.

### Patient Population and Baseline Clinical Variables

All patients in the regional GWTG-stroke database arriving by emergency medical service (EMS) to any of 8 CSC and 15 PSC hospitals in Chicago with 6 hours from the last time patient was confirmed to be symptom free (last known normal time) with a final diagnosis of sICH were included in this study. Patients that arrived at the hospital via private vehicle, transfer from another hospital, and any other mode of transportations were excluded. Patients with a final diagnosis of acute ischemic stroke, transient ischemic attack, subarachnoid hemorrhage, and stroke mimics were excluded. Clinical variables included were baseline demographic variables (sex, age, race), and comorbidities (e.g., hypertension, diabetes, atrial fibrillation). National Institute of Health Stroke Scale (NIHSS) at initial presentation, time of symptoms discovery, time of emergency department arrival, time of the initial CT scan were also captured from GWTG-Stroke. All data were entered by local site coordinators without central adjudication, interpretation or review, consistent with GWTG-Stroke data acquisition procedures.

### Outcomes

The primary outcomes were in-hospital mortality and favorable discharge disposition (defined as discharge to home or acute inpatient rehabilitation facility). Secondary outcomes included good neurologic outcome (defined as independent ambulatory status) at hospital discharge and stroke time metrics (symptoms-to-arrival-time, symptoms-to-CT-time, and door-to-CT-time), and inter-hospital transfer rates.

### Prehospital Stroke Transportation Protocol

The Chicago Stroke Advisory Subcommittee, in collaboration with the Chicago EMS system and Chicago Fire Department, implemented the 3-Item Stroke Scale (3I-SS) as a screening tool for identifying LVOs in patients presenting with suspected stroke within 6 hours of last known normal time. The 3I-SS scores range from 0 to 6 and assess three key parameters: level of consciousness, gaze, and motor function. Each parameter is assigned between 0 and 2 points, where 0 indicates normal function and 2 indicates severe impairment. Patients with suspected stroke as identified by the Cincinnati Prehospital Stroke Scale, were subsequently evaluated using the 3I-SS. Those who scored four or higher were triaged to a CSC, provided that the additional transport time did not exceed 15 minutes compared to transporting to the nearest PSC. This protocol was approved for implementation into the Chicago EMS system’s stroke transport policy in May 2018. The implementation included EMS education through both live and web-based training modules in September 2018, with the official go-live date set for November 28, 2018. All the participating hospitals were required to abstract data into GWTG-stroke database for quality improvement and monitoring and agreed to allow the American Heart Association Midwest Affiliate to aggregate data in a superuser account and share and report deidentified results with the Chicago stroke advisory subcommittee and EMS leadership monthly.

### Statistical Analysis

We compared data from April 2017 to August 2018 (17-month pre-implementation period) and from September 2018-Jan 2020 (16-month postimplementation period). Descriptive statistics were reported using proportions (95% confidence interval [CI]), medians (interquartile range [IQRs]) or mean (standard deviations [SDs]) when appropriate. Chi-square or Fisher exact were used for categorical data. Two-sided t test or Mann-Whitney test were used for comparison of mean or median of the continuous variables when appropriate.

We further used interrupted time series (ITS) for modeling each outcome, based on segmented linear regression which divides a time series into pre-implementation and post-implementation periods using September 2018 as the change point. This quasi-experimental technique is more rigorous than crude before and after comparisons, as it prevents confounding in the protocol effect estimation by accounting for temporal trends unrelated to the implantation. Hospital mortality rate, rate of good neurologic outcome and favorable discharge disposition at hospital discharge, median time from symptoms discovery to arrival and CT scan for each month were calculated. All models, one for each outcome, incorporate a baseline time trend, a protocol implementation indicator, and their interaction, while controlling for seasonal variations through quarterly indicators, stroke severity (NIHSS), and age, as well as for autoregressive moving-average model correlation structures where relevant. We also estimate the models using the official implementation date (November 28, 2018) as the protocol implementation date in separate sensitivity analyses. Change in level between the pre-implementation and post-implementation periods indicates step change, whereas a change in slope indicates a change in trend over time. All p values were 2-sided, and p <0.05 indicated statistical significance. All analyses were conducted using R (version 4.2.1).

## Results

### Baseline Characteristics

Of the total 15,929 patients screened in the Chicago regional GWTG-Stroke registry, 311 patients arrived at a hospital via EMS within 6 hours from the last known normal time and received a final diagnosis of sICH, where pre-implementation period included 111 patients and post-implementation period included 192 patients. Patients were excluded if they arrived by other means of transportation, has a diagnosis of ischemic stroke, transient ischemic attack, or stroke mimics (Figure 1). Included participants had a median age of 63 (IQR 53-74) years, 40% (n=125) were women, and 42% (n=130) were Black. NIHSS at initial presentation between pre- and post-implementation cohort were comparable (12 [5, 23] vs. 14 [6,22], p=0.9). Other baseline demographic characteristics and clinical features were listed in Table 1.

**Table 1.** Comparison of demographic and clinical characteristics of included spontaneous intracerebral hemorrhage patients before and after implementation of the prehospital stroke triage and transport policy.

| Characteristics | Study period April 2017-Jan 2020 |  |  |
| --- | --- | --- | --- |
|  | Pre-implementation (N=111) | Post-implementation (N=192) | p-value |
| Designation, n (%) |  |  | 0.01 |
| CSC | 25 (23%) | 71 (37%) |  |
| PSC | 86 (77%) | 121 (63%) |  |
| Age, median (IQR), years | 64 (55, 78) | 63 (52, 74) | 0.1 |
| Sex, n (%) |  |  |  |
| Female | 51 (46%) | 74 (39%) | 0.2 |
| Male | 60 (54%) | 118 (61%) |  |
| Race, n (%) |  |  | 0.2 |
| White | 45 (41%) | 58 (30%) |  |
| Black | 42 (38%) | 189 (51%) |  |
| Hispanic | 19 (17%) | 37(21%) |  |
| Asian | 11 (9.9%) | 16(8.3%) |  |
| Insurance status, n (%) |  |  | 0.4 |
| Medicare | 38 (46%) | 63 (41%) |  |
| Non-Medicare | 44 (54%) | 92 (59%) |  |
| NIHSS on arrival<sup>1</sup>, median (IQR) | 12 (5, 23) | 14 (6, 22) | 0.9 |
| Monthly External transfer (PSC to CSC) rate, median (IQR) | 0.85 (0.78, 0.88) | 0.76 (0.70, 0.83) | 0.081 |
| Symptoms discovery to arrival time, median (IQR), min | 86 (38, 355) | 65 (38, 278) | 0.4 |
| Door-to-CT time, median (IQR), min | 23 (10, 62) | 25 (13, 56) | 0.2 |
| Symptoms discovery to first CT, median (IQR), min | 131 (59,485) | 101 (63, 357) | 0.4 |
| In-hospital mortality, n (%) | 39 (14%) | 43 (12%) | 0.3 |
| Ambulation status at discharge, n (%)<sup>2</sup> |  |  | 0.5 |
| Ambulating independently | 11 (13%) | 20 (19%) |  |
| Unable to ambulate independently | 17 (21%) | 28 (26%) |  |
| Unable to Ambulate | 33 (40%) | 35 (32%) |  |
| Discharge disposition, n (%) |  |  | 0.8 |
| Home | 6 (5.4%) | 16 (8.3%) |  |
| Acute rehabilitation facility | 53 (48%) | 95 (49%) |  |
| Other healthcare facility<sup>3</sup> | 34 (31%) | 51 (27%) |  |
| Expired | 13 (12%) | 17 (9%) |  |

CSC: comprehensive stroke center; CT: computed tomography; IQR: interquartile range; NIHSS: national institute of health stroke scale; PSC: primary stroke center
<sup>1</sup>. Missing data: 31 for pre-implementation period, 56 for post implementation period.
<sup>2</sup>. Missing data: 29 for pre-implementation period, 84 for post implementation period.
<sup>3</sup>. Other Health Care Facility includes long term acute care hospital and skilled nursing facility.
Chi-square or Fisher exact were used for categorical data. Mann-Whitney test were used for comparison of median of the continuous variables. P values are two-sided, <0.05 is considered statistically significant.

**Figure 1.**
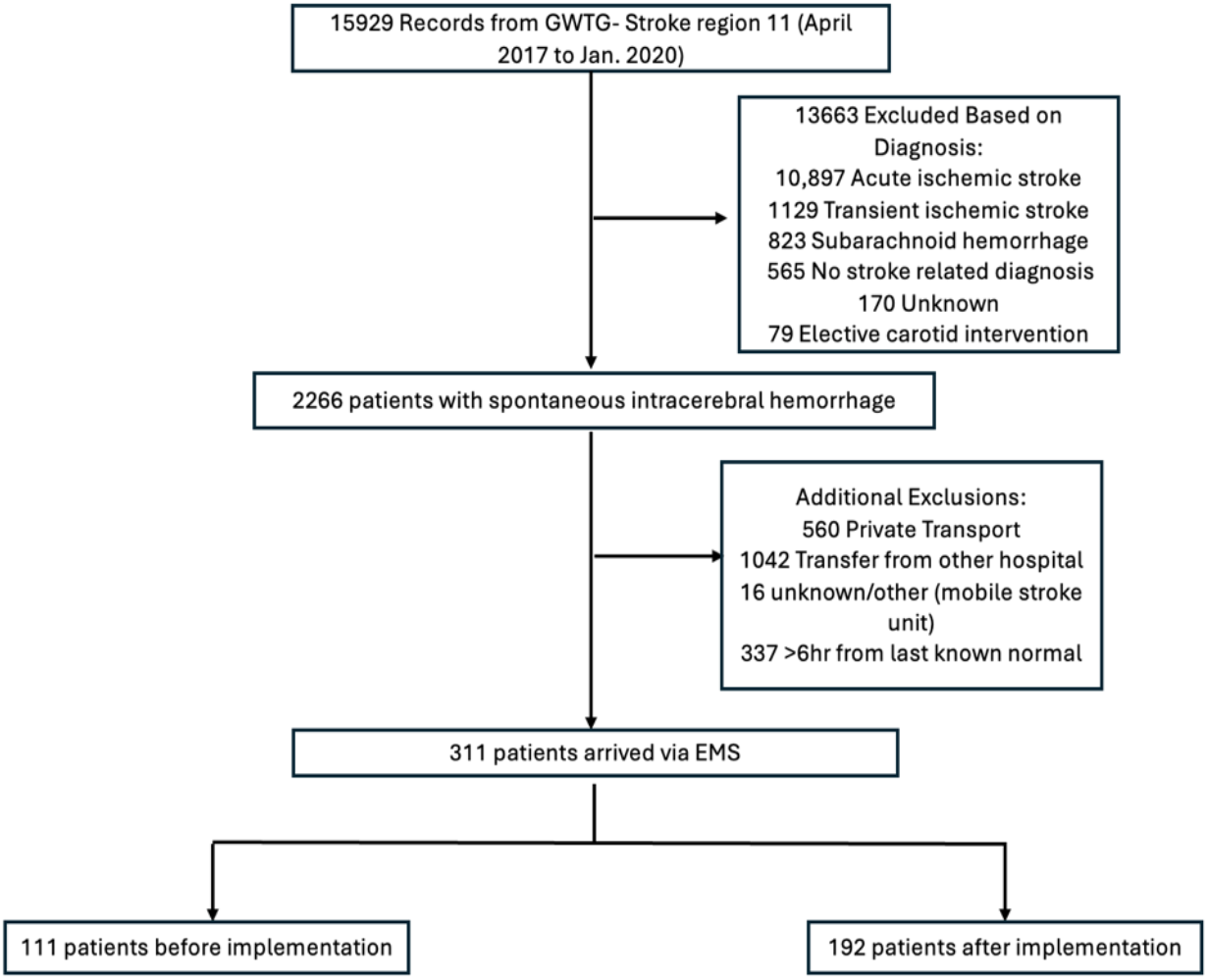
Study Cohort Assembly Flowchart.

### Primary Outcomes

For mortality, there was no difference in overall in-hospital mortality between pre- and post-implementation periods (12% vs. 9%, p=0.4) with ITS analysis confirming no significant immediate level (−5%, 95% CI: −31% to 21%, p=0.68) or trend (1%, 95% CI: −1% to 2%, p=0.34) change (Table 2). Similarly, favorable discharge disposition showed no change (58% to 64%, p=0.3) from pre-to post-implementation in unadjusted analysis and ITS analysis (level change: −20%, 95% CI: −77% to 38%, p=0.49; trend change: 1%, 95% CI: −2% to 4%, p=0.47; Table 2).

**Table 2.** Interrupted time series analysis of clinical outcomes at hospital discharge before and after implementation of the prehospital triage and transport policy among spontaneous intracerebral hemorrhage patients.

| Outcome | Variable | Effect estimates | 95% CI | p-value |
| --- | --- | --- | --- | --- |
| In-Hospital Mortality (%) | Level Change after Protocol Implementation | -0.05 | (-0.31, 0.21) | 0.68 |
|  | Post-Protocol Trend Change | 0.01 | (-0.01, 0.02) | 0.34 |
| Favorable Disposition at Discharge (Home/ACF; %) | Level Change after Protocol Implementation | -0.2 | (-0.77, 0.38) | 0.49 |
|  | Post-Protocol Trend Change | 0.01 | (-0.02, 0.04) | 0.47 |
| Favorable Ambulatory status at Discharge (%) | Level Change after Protocol Implementation | 0.11 | (-0.25, 0.48) | 0.53 |
|  | Post-Protocol Trend Change | -0.01 | (-0.03, 0.01) | 0.37 |

### Secondary Outcomes

For the secondary outcomes, good neurological outcome at discharge showed no significant difference between periods (13% vs 19%, p=0.4), with ITS analysis confirming no immediate level change (−13%, 95% CI: −54% to 29%, p=0.55) or trend change (0% per month, 95% CI: −2% to 1%, p=0.82; Table 2).

All time-based stroke metrics including door-to-CT time, symptom-to-arrival time and total symptom-to-CT time showed no significant difference between periods in unadjusted and ITS analysis (Table 1 and 3). Although CSC admission volume increased significantly post-implementation (37% vs. 23%, *p*=0.01; Table 1), this was not supported by ITS analysis (level change: −21%, 95% CI: −65% to 24%, *p*=0.35; trend change: 0% per month, 95% CI: −2% to 1%, *p*=0.82; Table 4). Additionally, there was no significant reduction in inter-hospital transfer rates from PSCs to CSCs (85% pre-vs. 76% post-implementation, *p*=0.081), consistent across both unadjusted and ITS analyses (Tables 1 and 3). Figure 2 summarized the impact of protocol implementation on all clinical outcomes.

**Table 3.** Interrupted time series analysis of operational and process outcomes before and after implementation of the prehospital triage and transport policy among spontaneous intracerebral hemorrhage patients.

| Outcome | Variable | Effect estimates | 95% CI | p-value |
| --- | --- | --- | --- | --- |
| Rate of Direct CSC Admission (% Total) | Level Change after Protocol Implementation | -0.21 | (-0.65, 0.24) | 0.35 |
|  | Post-Protocol Trend Change | 0.01 | (-0.01, 0.04) | 0.2 |
| Inter-hospital Transfer Rate (% Total) | Level Change after Protocol Implementation | 0.16 | (-0.18, 0.5) | 0.37 |
|  | Post-Protocol Trend Change | -0.01 | (-0.03, 0) | 0.16 |
| Symptoms - Arrival (minutes) | Level Relative Change after Protocol Implementation (proportion of pre-protocol average) | 0.97 | (0.3, 3.11) | 0.96 |
|  | Post-Protocol Trend Change | 1.02 | (0.96, 1.08) | 0.49 |
| Arrival - First CT (minutes) | Level Relative Change after Protocol Implementation (proportion of pre-protocol average) | 1.55 | (0.55, 4.38) | 0.41 |
|  | Post-Protocol Trend Change | 0.99 | (0.93, 1.04) | 0.59 |
| Symptoms - First CT (minutes) | Level Relative Change after Protocol Implementation (proportion of pre-protocol average) | 0.92 | (0.44, 1.95) | 0.83 |
|  | Post-Protocol Trend Change | 1.01 | (0.97, 1.04) | 0.74 |

CI: confidence interval; CSC: Comprehensive stroke center; CT: Computed Tomography

**Figure 2.**
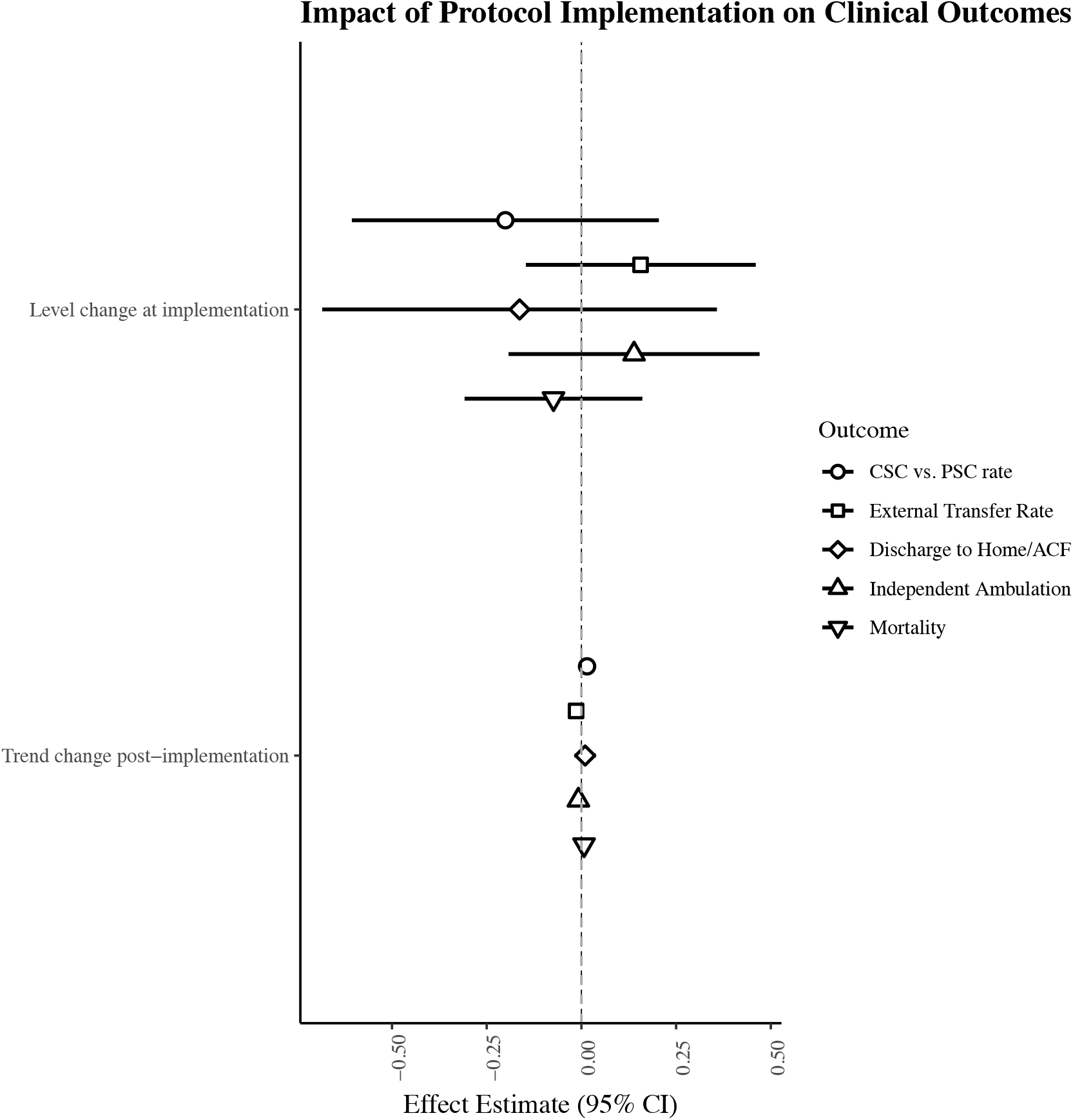
Impact of LVO-Focused Prehospital Transport Protocol Implementation on Clinical Outcomes in Spontaneous ICH: Forest Plot of ITS Estimates

### Sensitivity Analysis

A sensitivity analysis was conducted using the official protocol implementation date of November 28, 2018, instead of the initial rollout in September 2018. Supplemental Table 1 compares baseline demographics using the November cut-off. This analysis demonstrated a significant reduction in monthly inter-hospital transfer rates from PSCs to CSCs (85% to 74%, *p*=0.005, Supplemental Table 1), though ITS analysis revealed no significant immediate level change (−7%, 95% CI: −49% to 35%, *p*=0.74) or trend change (−1%, 95% CI: −2% to 1%, *p*=0.54) (Supplemental Table 2). There was also a trend toward increased discharge to a favorable location post-implementation (53%, 95% CI: 42–56% vs. 57%, 95% CI: 52–65%, *p*=0.051), but ITS analysis again showed no immediate level change (1%, 95% CI: −71% to 71%, *p*=0.98) or trend change (1%, 95% CI: −2% to 4%, *p*=0.67). No significant differences in mortality, functional outcomes, or time-based metrics were observed using the November 2018 cut-off (Supplemental Table 2).

## Discussion

Our study evaluated the impact of a regional LVO-focused prehospital stroke triage protocol on sICH in an urban stroke system of care. Although the protocol increased the proportion of sICH admissions to CSCs, this shift was not supported by ITS and did not translate to improved clinical or process outcomes for sICH patients. Our analysis revealed no significant differences in good neurological outcomes, mortality rates, or critical time-metrics including door-to-CT and symptom-to-CT times following protocol implementation. While there was a small trend of increased favorable discharge disposition in the sensitivity analysis, this was not corroborated by the ITS analysis, suggesting a broader temporal trend rather than a direct effect of the policy.

Our findings align with prior studies that have evaluated the impact of direct CSC transport for sICH patients. The secondary analysis of the RACECAT trial ^9^ found that sICH patients transported directly to CSCs from non-urban settings did not experience improved outcomes and, in some cases, had worse functional recovery, potentially due to prehospital transport delays. Furthermore, studies have also shown that earlier admission to CSCs and faster diagnosis of sICH via mobile stroke units despite facilitating more rapid systolic blood pressure reduction and shorter dispatch-to-imaging-times did not translate into improved functional outcome.^11,12^ Another analysis of the GWTG-Stroke registry demonstrated that rapid stabilization and early medical management at the closest hospital are preferred strategies for sICH care, particularly in urban settings with short transport distances.^13^ One possible explanation for these findings is that sICH management primarily relies on early blood pressure control, airway, breathing, circulation optimization, and reversal of anticoagulation when applicable, which can be initiated at both PSCs and CSCs. Unlike EVT in LVO patients, where treatment delays significantly impact outcomes,^6–8^ sICH patients may not derive the same magnitude of benefit from bypassing the closest PSC. Our results corroborate these prior studies in showing that a centralized triage approach does not necessarily benefit sICH patients, though importantly, we did not find evidence of harm from the protocol.

Our findings also highlight critical limitations of current LVO-focused prehospital tools for sICH triage. While scales like 3I-SS effectively identify anterior circulation LVOs and may identify severe strokes including lobar ICH, they were not developed for ICH detection in the field. This underscores the need for more nuanced hemorrhage-specific screening tools that account for its unique pathophysiology and management priorities to improve field identification of sICH patients who might benefit from direct CSC transport. Recent advances in ICH detection scales showed particular promise. Freixa-Cruz et al. developed a predictive model (AUC=0.82) using headache, GCS<8, SBP>160 mmHg, and male sex to differentiate ICH from LVO, while the Japan Urgent Stroke Triage score achieved even higher accuracy (AUC=0.84) for stroke subtype classification.^18,19^ Implementation of such validated, ICH-specific scales could potentially improve outcomes through timely and appropriate triage and interventions. With the increasing adoption of MIS following the ENRICH trial, there is renewed interest in optimizing early access to MIS-capable CSC. MIS offers the potential to reduce hematoma volume and improved functional outcomes, particularly when performed early—ideally within 24 hours of symptom onset.^146^ As such, timely identification of surgical candidates and direct CSC transport may have greater utility in the future. However, during our study period, MIS utilization was still uncommon, limiting our ability to assess its impact on patient outcomes. Future protocols should prioritize the prospective validation of hemorrhage-specific triage tools across diverse EMS systems to better identify candidates for early surgical intervention and evaluate transport strategies that balance rapid stabilization at PSC against timely access to advanced surgical care at MIS-capable CSCs. Additionally, studies evaluating the long-term functional outcomes of sICH patients under different triage models are also needed to better understand the relationship between transport decisions and long-term functional outcomes.

Our study has several limitations. First, it is a single-region study in a highly urban setting with short transport distances, which may not generalize to rural or suburban areas where CSC access is more limited. Second, the retrospective and observational nature of our study introduces the potential for residual confounding, despite the use of rigorous statistical methods such as ITS analysis. Third, reliance on the GWTG-Stroke database may have limited our ability to capture certain clinical details, such as variations in prehospital blood pressure management or hospital-level sICH protocols and key intermediary outcomes, such as time to blood pressure control or anticoagulant reversal, time to neurosurgical consultation, and escalation of care decisions. Forth, lack of long-term functional outcome data limits our ability to assess the full impact of the protocol on sICH patients’ recovery. Indeed, recovery from sICH may be best measured at 180-or 365-days post-event.^15–17^ Lastly, while adequately powered to detect large effect sizes, the sample size may have been insufficient to rule out potentially clinically important differences reflected in the wide 95% confidence interval for the primary outcome (−31% to 21%).

## Conclusion

While the implementation of a regional prehospital stroke transport protocol designed to triage patients with suspected LVO preferentially to CSCs significantly alter transport patterns for sICH patients, there was no impact on clinical outcomes or secondary process outcomes. These findings underscore the need for prehospital tools and transport protocols specifically tailored to ICH rather than repurposing ischemic stroke triage tools. Future studies should focus on refining prehospital identification methods and evaluating the evolving role of direct CSC transport in the context of emerging minimal invasive surgical and neurocritical care interventions.

## Data Availability

Data used in this manuscript is available upon request.

